# Lived Experiences of Registered Nurses Delivering Care During Winter Conditions: An Interpretative Phenomenological Study

**DOI:** 10.64898/2025.12.02.25341229

**Authors:** Fernan Torreno

**Affiliations:** Alhada Armed Forces Hospital Taif KSA

**Keywords:** Community nursing, Continuity of care, Lived experiences, Patient safety, Phenomenology, Qualitative research, Resilience, Telehealth, Winter healthcare delivery, Workforce planning

## Abstract

**Background:** Winter conditions intensify operational demands on healthcare services, particularly in community and rural settings where nurses often travel to deliver care. Environmental challenges, including hazardous road conditions and limited accessibility, can affect nurse well-being and patient safety. Few studies have explored how nurses personally experience and adapt to winter-related care delivery constraints.

**Objectives:** To explore the lived experiences of registered nurses practicing during winter conditions, identify key challenges, examine adaptive strategies, and inform workforce and policy development to strengthen seasonal preparedness.

**Methods:** A qualitative study using interpretative phenomenological analysis (IPA) was conducted with 20 registered nurses. Participants were selected through purposive sampling based on active practice and minimum one winter season of care delivery. Semi-structured interviews for one hour were transcribed and thematically analyzed. Study rigor was supported by reflexive journaling, peer debriefing, and thick descriptive reporting.

**Results:** Four core themes emerged: (1) physical strain and cold exposure, (2) psychosocial stress and resilience, (3) patient safety risks and care disruption, and (4) adaptive strategies and professional innovation. While winter significantly increases workload and risk, participants demonstrated strong resilience, frequently using flexible scheduling, telehealth, and collaborative decision-making to maintain continuity of care.

**Conclusion:** Winter conditions considerably challenge nursing practice but simultaneously promote adaptability and teamwork. System-level support is required to shift from reactive to proactive winter preparedness. Policy recommendations include winter-specific training, improved access to protective resources, telehealth infrastructure investment, and resilience-focused workforce strategies.

**Policy Implications:** Enhancing winter preparedness is essential to protect nurse well-being and maintain care continuity during adverse environmental conditions.

## Introduction

Healthcare delivery is profoundly shaped by environmental conditions, with winter recognized as one of the most challenging periods for frontline healthcare professionals. Nurses, particularly those working in community health, home care, and rural outreach settings, often provide essential services under harsh conditions that include hazardous travel routes, reduced visibility, temperature extremes, and irregular access to medical resources^1^. These winter-related obstacles increase operational risk and can directly threaten continuity of care, especially for patients with chronic illnesses or limited mobility^2^.

Seasonal fluctuations further contribute to increased workload pressures. Winter is consistently associated with higher rates of hospital admission due to exacerbations of respiratory and cardiovascular conditions, increased communicable disease transmission, and weather-related physical injuries^3^. In rural and remote regions, where healthcare infrastructure may already be limited, accessibility barriers become more pronounced□. Existing studies have highlighted systemic strain during winter, yet few have examined these challenges from the perspective of nurses themselves, whose frontline role is essential in sustaining care delivery□.

Understanding nurses’ lived experiences is critical, as it allows for deeper insight into the human, operational, and ethical dimensions of seasonal care provision. Resilience has been identified as a core component underpinning nurses’ ability to cope with adversity, strengthened by peer collaboration and shared decision-making□. Innovative adaptations, such as telehealth implementation, have emerged as viable alternatives to in-person care when travel becomes unsafe, particularly following the digital transformation accelerated by the COVID-19 pandemic□. Despite such positive developments, winter-specific support strategies—such as safety protocols, adaptable scheduling, and targeted training—remain inconsistently integrated across healthcare systems□.

Consequently, understanding how nurses navigate winter-related challenges is vital for informing health policy development, resource planning, and workforce resilience strategies. This study addresses an important gap by exploring the lived experiences of registered nurses delivering care during winter conditions. It aims to provide evidence-based recommendations to support sustainable practice and improve care continuity during adverse seasonal periods.

Specifically, this study seeks to:

1. Examine the physical, psychosocial, and environmental challenges faced by nurses during winter practice^1^;
2. Identify coping strategies and resilience mechanisms that enable continuity of care^2^;
3. Assess the impact of winter working conditions on nurse well-being and professional responsibility^3^; and
4. Recommend winter-specific interventions at workforce and policy levels to enhance practice sustainability□.

This research contributes to growing discourse on climate-related healthcare challenges and emphasizes the urgency of integrating winter preparedness into nursing practice, health system design, and strategic workforce planning.

## Methods

### Study Design

An interpretative phenomenological analysis (IPA) qualitative research design was adopted to gain deep insight into the lived experiences of registered nurses engaged in winter practice. IPA is acknowledged for its sensitivity to the personal and contextual interpretation of experiences, making it particularly suitable for exploring complex occupational challenges in nursing^1^. This approach facilitated rich, nuanced interpretation rather than mere description, aligning with the aim of understanding how nurses construct meaning from their encounters with harsh winter conditions.

### Research Setting and Participants

This study was conducted across community health, home care, rural outreach, and public health settings situated within areas known for consistent winter severity. Purposive sampling was used to identify nurses with direct, relevant experience, ensuring depth and richness of data^2^. Registered nurses were eligible if they (a) were actively practicing within the last three years, (b) had completed at least one full winter season involving patient care, and (c) were proficient in English. A total of 20 participants were recruited, representing both urban (n=12) and rural (n=8) regions, with professional experience ranging from 2 to 25 years. Nurses in non-direct care roles, trainees, and those without winter exposure were excluded to maintain relevance and interpretability.

### Data Collection Procedures

Data were collected via one-on-one semi-structured interviews conducted between [insert months/year]. Interviews were held either in person or via secure videoconferencing platforms, depending on location accessibility and participant preference. Each session lasted between 60 and 90 minutes. An interview guide, developed based on existing literature and refined through pilot testing, explored themes such as environmental barriers, psychological responses, patient safety concerns, workload pressures, and adaptation strategies^3^. All interviews were audio recorded with participant consent and transcribed verbatim. Field notes were maintained during and immediately following interviews to document non-verbal cues, contextual details, and researcher reflections.

### Data Analysis Approach

Analysis was conducted using IPA following the steps described by Smith et al.^1^. The process involved: (1) immersive reading of transcripts to develop contextual familiarity; (2) initial line-by-line coding to identify meaningful statements; (3) grouping codes into emergent themes; (4) synthesizing themes into superordinate conceptual categories; and (5) interpretative integration with supporting literature□. Coding was managed using NVivo software. To enhance credibility, 20% of interviews were independently coded by a second researcher, and iterative discussions were held to refine theme development and resolve discrepancies.

### Reflexivity and Researcher Positionality

Reflexivity was systematically integrated throughout the project to mitigate interpretive bias. The lead researcher maintained a reflective journal documenting personal assumptions, emotional responses, and interpretative decisions during data collection and analysis□. Peer debriefing sessions conducted with subject matter experts provided an additional layer of scrutiny and supported analytic consistency□.

### Ensuring Study Rigor

Trustworthiness was maintained using Lincoln and Guba’s framework, including credibility, transferability, dependability, and confirmability. Techniques for enhancing rigor included thick descriptive data presentation, triangulation through cross-verification of coded themes, maintenance of an audit trail, and context-specific participant representation□. Although direct member checking was not feasible due to geographic dispersion, transcript validation was undertaken to confirm narrative accuracy.

### Ethical Considerations

Ethical approval was granted by the Region 1 Ethical Review Committee (Reference: NUR2025-DD826A). All participants provided informed written consent and were advised of their voluntary participation and right to withdraw at any time without consequence□. Confidentiality was ensured through the assignment of pseudonyms and secure handling of all research materials.

### AI Use Disclosure

Artificial intelligence was employed solely for the refinement of manuscript structure and linguistic clarity. All conceptual development, interview facilitation, data analysis, and interpretative processes were conducted independently by the research team to preserve authenticity and academic integrity.

## Results

Analysis of interview data revealed four interconnected themes describing nurses’ lived experiences during winter practice: (1) Physical Strain and Environmental Exposure^1^, (2) Psychosocial Stress and Resilience^2^, (3) Patient Safety Risks and Care Delivery Challenges^3^, and (4) Adaptive Strategies and Professional Innovation□. Although participants reported significant strain, many expressed that winter practice fostered resourcefulness and strengthened professional identity.

### Participant Characteristics

A total of 20 nurses contributed to the study. Table 1 outlines demographic characteristics.

**Table 1.** Participant Demographics (n = 20)

| Attribute | Category | n (%) |
| --- | --- | --- |
| Gender | Female | 15 (75%) |
|  | Male | 5 (25%) |
| Region | Urban | 12 (60%) |
|  | Rural | 8 (40%) |
| Experience | 2–5 years | 4 (20%) |
|  | 6–10 years | 6 (30%) |
|  | 11–20 years | 7 (35%) |
|  | 21+ years | 3 (15%) |
| Practice Setting | Community | 7 (35%) |
|  | Home care | 5 (25%) |
|  | Rural outreach | 4 (20%) |
|  | Public health | 4 (20%) |

Theme 1: Physical Strain and Cold Exposure^1^

Participants described the physical burden of practicing in extreme weather conditions. Extended travel on icy roads and working in low-temperature settings contributed to fatigue and musculoskeletal discomfort. One nurse explained:

***“By the time I arrive at my second visit, I’m already exhausted—not from the work itself, but because moving around in multilayered clothes takes so much effort.” (P4)***

Common physical effects included reduced mobility, increased energy exertion, and heightened risk of illness. Nurses also emphasized limited access to heated spaces during home visits.

Theme 2: Psychosocial Stress and Resilience^2^

Emotional strain was reported across all participants, with rural nurses noting increased feelings of isolation and anxiety about safety. However, resilience was often reinforced by teamwork:

***“Driving alone in snowstorms was mentally draining, but regular check-ins with colleagues helped me cope.” (P11)***

Figure 1 summarizes reported emotional impact.

**Figure 1.**
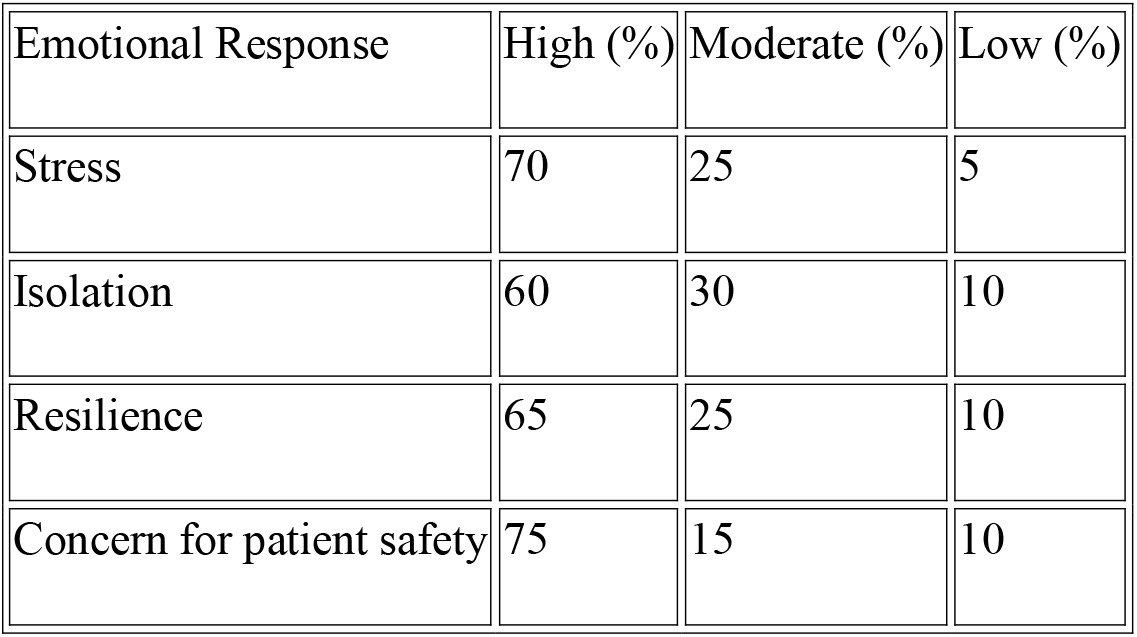
Reported Emotional Responses (n = 20)

Theme 3: Patient Safety Risks and Care Delivery Challenges^3^

Winter conditions disrupted care delivery due to transport restrictions, road closures, and delays in emergency support. Participants expressed concern for vulnerable patients:

***“When we can’t get there on time, especially for elderly clients, the consequences can be serious.” (P7)***

Care prioritization emerged as a common strategy, with acute cases scheduled earlier in the day to avoid worsening weather.

Theme 4: Adaptive Strategies and Innovation□

Participants described several adaptive approaches that enabled continuity of care. Table 2 highlights the most frequently cited strategies.

**Table 2.** Winter Adaptation Strategies.

| Strategy | n (%) |
| --- | --- |
| Layered thermal clothing | 18 (90%) |
| Flexible scheduling | 17 (85%) |
| Use of telehealth consultations □ | 14 (70%) |
| Collaborative planning | 19 (95%) |
| Vehicle safety equipment | 15 (75%) |

***“Using telehealth was a game changer on days we just couldn’t reach patients—it kept care consistent even in extreme weather.” (P5)***

### Summary of Findings

While winter practice significantly increases physical and emotional strain among nurses, it simultaneously promotes resilience, professional collaboration, and innovative use of care delivery models. These findings inform the need for system-supported winter preparedness strategies.

## Discussion

This study explored the lived experiences of registered nurses delivering care during winter conditions, revealing substantial challenges related to physical strain, emotional impact, patient safety risks, and adaptive practice responses. Findings indicate that winter significantly intensifies workload demands and logistical complexity, particularly for nurses working in community and rural settings, where travel hazards, limited resources, and isolation are most prevalent^1^. These findings align with previous research identifying those nurses practicing in challenging environments experience heightened strain, especially when working in mobile or distributed care roles^2^.

### Physical impact and occupational strain

were dominant concerns, with participants frequently referencing fatigue related to cold exposure, wearing heavy clothing, and navigating slippery surfaces during patient travel. Similar findings have been observed in research describing environmental strain among community nurses^3^. The physiological stress reported supports the need for winter-specific occupational health planning, including provision of thermal protection, ergonomic equipment, and safe travel protocols for healthcare workers.

### Psychosocial stress was also prominent

particularly among rural practitioners, who reported feelings of isolation and vulnerability during winter travel. However, participants emphasized the role of peer support and team-based collaboration in mitigating emotional strain, consistent with resilience studies suggesting that social connectedness is foundational to adaptive coping in professional nursing practice□. This highlights the importance of workplace cultures that encourage relational safety and shared accountability during high-pressure periods.

A critical component of winter practice relates to **patient safety and continuity of care**. Nurses reported ethical dilemmas when weather conditions delayed or prevented access to vulnerable patients. Previous studies note similar challenges in remote healthcare settings, where infrastructure limitations can exacerbate health inequities□. Although nurses often used early appointments or route prioritization to ensure access to at-risk patients, many expressed concern about increased risk of harm and emergency escalation during severe weather. Findings underscore the need for system-level intervention, such as contingency transport provisions and mobile emergency planning resources.

### Adaptive behaviors and innovation emerged as a major protective theme

reflecting professional creativity in response to environmental limitations. Nurses frequently implemented flexible scheduling, with high adoption of telehealth as an alternative when travel was unsafe. This supports existing evidence of telehealth’s efficacy in community care delivery□. However, participants emphasized that its success was dependent on digital access and organizational support. This suggests that telehealth integration should form part of winter preparedness modeling rather than being used reactively. Collaborative decision-making and pre-planning were reported as central to winter resilience, reinforcing literature calling for proactive rather than emergency-based adaptation strategies□.

While results indicate strong individual resilience among nurses, the study also reveals a dependency on personal adaptation due to insufficient structural support. Participants described repeated reliance on self-directed strategies rather than formal protocols or resourced initiatives. This corresponds with previous findings suggesting nurse resilience may mask the absence of system-level planning□. There is growing recognition that relying solely on personal coping undermines long-term workforce sustainability. Consequently, resilience should be reframed as a collective and systemic principle supported by policy, infrastructure, and leadership-led strategies.

#### Study Limitations

The study was geographically constrained to regions with defined winter profiles, which may limit transferability to areas with milder climates. Data were self-reported and subject to recall bias. Member checking was not completed due to logistical limitations, although transcript validation enhanced credibility. Given the qualitative nature of the study, findings are not generalizable but are transferable to practice contexts with comparable profiles.

Future research should examine winter nursing experiences across broader geographic regions, evaluate the efficacy of winter preparedness policies, and assess the impact of climate variability. Simulation-based winter response training and longitudinal evaluation of telehealth adoption may offer valuable insight into scalable solutions.

## Conclusion

Winter conditions place substantial physical, logistical, and emotional strain on nurses, yet simultaneously foster professional resilience and innovation. Participants demonstrated a strong capacity for adaptation, utilizing strategies such as collaborative planning, flexible scheduling, and telehealth implementation. However, findings suggest that these adaptations are often self-directed responses to system limitations, rather than evidence of effective institutional planning.

To strengthen winter practice in nursing and safeguard patient outcomes, healthcare systems must transition from reactive to proactive models of preparedness. This includes integrating winter-specific training, provision of protective gear, expanding telehealth infrastructure, and ensuring safe transport access for community-based practitioners. Workforce resilience must be addressed not only as an individual attribute but as a supported organizational framework that promotes psychological safety, collaborative care decision-making, and accountability sharing.

The study provides timely evidence that winter-related challenges are not solely operational, but also occupational health concerns that impact long-term workforce sustainability. As environmental variability continues to affect care delivery, policy reform targeting seasonal preparedness is essential. Implementation of winter-specific resilience and safety protocols can help safeguard nurse well-being, protect vulnerable patient groups, and enhance continuity of care.

In summary, the lived experiences of nurses demonstrate the duality of winter practice: while significantly increasing occupational strain, it also provides opportunities to advance resilience-driven professional development. Strengthening system-level frameworks to support this resilience is key to ensuring sustainable care provision under adverse environmental conditions.

## Data Availability

Availability of Data and Materials
The de-identified transcripts and coding framework used in the study are available from the corresponding author upon reasonable request and subject to ethical approval requirements.

https://data.mendeley.com/drafts/2dwy33fjch

## Declarations

Ethics Approval and Consent to Participate

## Ethical Considerations

This study received ethical approval from the **Region I Ethical Review Committee, Department of Health – Center for Health Development, San Fernando City, La Union, Philippines**. The Committee reviewed the study protocol (Reference: NUR2025-DD826A) and granted full approval. All participants provided informed written consent prior to participation. They were informed of their voluntary participation, their right to withdraw at any time without consequence, and the measures taken to ensure confidentiality. Confidentiality was maintained through the assignment of pseudonyms and secure handling of all research materials. No waiver of ethical approval was sought or granted; full approval was obtained.

### Consent for Publication

Not applicable. The manuscript contains no identifiable personal data, images, or direct reference to individuals. All interview excerpts were anonymized.

### Availability of Data and Materials

The de-identified transcripts and coding framework used in the study are available from the corresponding author upon reasonable request and subject to ethical approval requirements.

### Funding

No external funding was secured for this study. The research was independently conducted and self-funded by the principal investigator.

### Authors’ Contributions

Dr. **Fernan Torreno** conceptualized the study, conducted participant interviews and data analysis, and developed the manuscript. All authors contributed to the editing process and approved the final version of the paper.

## Acknowledgements

The author thank the registered nurses who generously shared their time and experiences. Appreciation is extended to professional networks that assisted with recruitment and to colleagues who provided reflexive feedback during analysis.

## AI Disclosure

Artificial Intelligence was used only for **language refinement, structure improvement, and formatting assistance** during manuscript preparation. It was **not involved in study design, data collection, coding, or interpretation**. All core research activities and final analysis were conducted solely by the research team, in accordance with ethical research standards.

